# Disability, Chronic Pain, and Opioid Use Disorder Treatment Disparities: A National Cohort Analysis from the *All of Us* Research Program

**DOI:** 10.64898/2026.08.09.26359632

**Authors:** Jessica Williams, Bailey W. Osweiler, Junlachak P. Siriprakorn, Phillip L. Marotta

**Author notes:** **Corresponding Author:** Jessica Williams, MD Contact; *Permanent Address:* 660 South Euclid Ave. MSC-8134-17-04, Saint Louis, MO 63110, USA. **Previous Presentation:** This work was previously presented in an oral session at the 2026 College on Problems of Drug Dependence (CPDD) Annual Meeting on 6/15/2026. The abstract is included in the meeting agenda for attendees. **Declarations of interest:** None. **Author Contributions:** All authors had full access to all the data in the study and take responsibility for the integrity of the data and the accuracy of the data analysis. **CRediT Author Statement:** The following roles are reported using the CRediT (Contributor Roles Taxonomy). **Jessica Williams:** Conceptualization, Methodology, Investigation, Writing – original draft, Writing – review & editing, Project administration. **Bailey W. Osweiler:** Conceptualization, Methodology, Investigation, Software, Data curation, Formal analysis, Validation, Visualization, Writing – original draft, Writing – review & editing. **Junlachak P. Siriprakorn:** Conceptualization, Methodology, Investigation, Validation, Visualization, Writing – review & editing. **Phillip L. Marotta:** Conceptualization, Writing – review & editing, Supervision. **Role of the Funder/Sponsor:** N/A. **Disclaimers:** None to report. **Conflicts of Interest:** None to report.

## Abstract

**Background:** People with disabilities (PWD) represent over one-quarter of the US population and disproportionately experience chronic pain, yet limited research explores disparities they face in opioid use disorder (OUD) treatment.

**Objective:** To examine disparities across disability status regarding opioid use disorder (OUD)-related outcomes and understand how chronic pain interacts with these associations.

**Methods:** We completed a cross-sectional, secondary analysis of data from the All of Us Research Program, including 370,722 adults with electronic health record data available between January 2021-September 2023. We identified prevalence of disability, chronic pain, OUD, receipt of medications for OUD (MOUD), and OUD remission using diagnostic codes. We performed interaction analyses between chronic pain, disability subtype, and MOUD receipt in affecting OUD outcomes.

**Results:** OUD was more common among individuals with physical (aOR: 2.74, 95% CI: 2.54-2.95), cognitive (2.19, 1.94-2.45), and multiple disabilities (2.43, 2.19-2.68), compared to those without disabilities. Among patients with OUD, those with physical disabilities were less likely to receive MOUD (0.81, 0.69-0.94). Compared to those without disabilities, chronic pain was associated with higher probabilities of OUD diagnosis and lower probabilities of MOUD and OUD remission across all subjects. These relationships were stronger for OUD diagnosis in cognitive disabilities, MOUD in multiple disabilities, and OUD remission in physical disabilities.

**Conclusions:** Disability and chronic pain jointly shape disparities in OUD treatment and underscore the urgent need for care models that integrate OUD treatment with pain management and address the unique access challenges faced by people with disabilities.

## 1. Introduction

There has been extensive research centered on the U.S. opioid crisis, yet investigations at the intersection of disability, chronic pain, and opioid use disorder (OUD) remain sparse. Although disability affects 28.7% of U.S. adults,^1^ we know little about how people with disabilities (PWD) navigate the opioid use care cascade, from diagnosis to treatment initiation and sustainment.^2^ Existing knowledge in this domain is drawn almost entirely from secondary analyses of the National Survey on Drug Use and Health (NSDUH)^3^ and insurance claims^4^— datasets that were not designed to comprehensively assess disability status or chronic pain. What we can glean from NSDUH data is troubling: PWD, who experience a range of limitations in hearing, vision, cognition, mobility, self-care, and independent living,^1^ have disproportionately high rates of opioid misuse, OUD, and related morbidity and mortality, especially when affected by pain.^4–6^ Worse, PWD among Washington Medicaid enrollees were 40% less likely to receive any medications for OUD (MOUD) compared to people without disabilities (PWOD).^2^ Further, there does not appear to be existing literature regarding PWD’s likelihood of achieving and sustaining remission from OUD.

OUD can be further compounded by the presence of chronic pain, which affects 52.4% of PWD, a rate triple that of PWOD.^7^ Chronic pain may function as a key pathway through which PWD are more likely to be exposed to opioids and develop OUD.^8^ Medicaid data indicate that comorbid disability and chronic pain are associated with elevated risk of OUD and overdose, likely reflecting greater exposure to long-term prescribing for pain management, limited access to nonpharmacologic pain treatments, and fragmented specialty care.^9^ MOUD are known to effectively reduce relapse, overdose, and death from opioid use,^10^ and can help alleviate chronic pain themselves^11^; though PWD encounter substantial barriers to initiating and sustaining treatment.^2,12,13^ Even if OUD treatment centers were more accessible, they often do not manage chronic pain, which is frequently reported as a primary factor in OUD relapse.^14^ High prevalence of chronic pain among PWD may complicate treatment decision-making and care coordination, though empirical evidence on pain’s role in MOUD receipt and recovery outcomes is limited.

This study directly addresses these gaps by using the *All of Us* Research Program’s diverse national cohort data to investigate how disability and chronic pain interact with regard to OUD diagnosis, MOUD receipt, and remission. We hypothesized that PWD would exhibit higher odds of OUD diagnosis, lower rates of MOUD receipt, and reduced OUD remission rates compared to PWOD. Furthermore, we expected the presence of chronic pain would worsen these disparities. We anticipated that specific disability subtypes, especially physical disabilities and those with multiple disability subtypes, would experience the most pronounced disadvantages when chronic pain is also present. By clarifying where disparities emerge and identifying how chronic pain can modify treatment patterns and remission outcomes, this work aimed to provide urgently needed evidence to inform more equitable and accessible addiction care. Addressing the intersection of disability, chronic pain, and OUD is essential for ensuring public health strategies and clinical interventions serve all populations equitably rather than reinforce existing disparities.

## 2. Methods

### 2.1. Data source and inclusion criteria

The *All of Us* research program collected data from voluntary participants at academic, Veterans Affairs, and community health centers nationwide, with emphasis on recruiting a diverse participant population, including those underrepresented in biomedical research (e.g., PWD, populations with reduced healthcare access).^15^ For the present study, we accessed the *All of Us* Registered Tier Dataset v8, including individual-level electronic health record (EHR), wearable, physical measurement, and survey data. We included participants with EHR data available between January 2021-September 2023 and who were adults (≥18 years) at the beginning of the study period. This study was exempted from review by the [blinded for review] Institutional Review Board (####). STROBE reporting guidelines for cross-sectional studies were followed.

### 2.2. Measures

#### 2.2.1. Primary outcome: OUD diagnosis

A binary indicator was created of whether a participant had 1+ OUD diagnosis, including ICD9 codes 304.0, 304.7, 305.5, and ICD10 codes under F11.^16^

#### 2.2.2. Secondary outcomes

*MOUD receipt, OUD remission.* Binary indicators were created for 1+ MOUD prescriptions and 1+ diagnosis of OUD in remission. MOUD included prescriptions for methadone, buprenorphine, or naltrexone (generic and trade names), and excluded Buprenex and Butrans, as these are primarily used for chronic pain.^17^ OUD remission included ICD9 codes 304.03, 304.73, and 305.53 and ICD10 “F11” codes with the last digit equal to x1.^16^

#### 2.2.3. Independent variable: disability diagnoses categorized into subtypes

The primary predictor of interest was a binary indicator of disability and a categorical indicator of disability subtype. Because no standardized approach exists for identifying disability using clinical or administrative data, we operationalized disability using a multidimensional, claims-based classification^18^ that incorporates ICD-9, ICD-10, CPT, and HCPCS codes (see **Supplemental Table 1**), consistent with prior studies.^19,20^ We manually classified disabilities into categories: “cognitive,” “sensory,” or “physical.” To accommodate individuals with indicators of more than one disability subtype, we created an additional “multiple” disability subtype.^21^ Those with multiple disability subtypes were not also counted in the individual subtype group counts.

#### 2.2.4. Covariates

Demographics (race, sex, gender, age at beginning of study period) were included as covariates. As pain is related to both disability and OUD, a chronic pain covariate was included via ICD9 codes Z79.891, 338.2, 338.4, and ICD10 codes G89.2, G89.4.^22,23^

### 2.3. Statistical Analysis

Descriptive analysis characterized demographic and clinical characteristics by disability subtype. Univariate logistic regression examined relationships between disability and OUD, MOUD, or remission, while multivariable analyses adjusted for sociodemographic covariates (gender, age, race, and ethnicity) and chronic pain. The OUD remission model additionally controlled for MOUD. Adjusted odds ratios (aOR) and 95% confidence intervals (CI) were reported. To test for potential effect modification, we included interaction terms between disability status (ref: no disability) and chronic pain (ref: no pain),^24^ as well as between disability status and MOUD receipt. We reported significant regression interaction results as predicted probabilities using marginal standardization, controlling for mean age and weighted averages across categorical variables.^25^ Sensitivity analysis tested for pairwise absolute differences in predicted probabilities of remission within each disability subtype by MOUD and chronic pain using the ggeffects “test_predictions” function.^26^ Significance for all tests was assessed at a two-sided α=0.05. We used a Bonferroni correction to control for Type I error inflation when performing multiple pairwise comparisons (α=0.01).^27^

The cohort was identified using the All of Us Researcher Workbench, where we searched for inclusion criteria codes. Because exposures, outcomes, and the chronic pain covariate were defined by the presence or absence of qualifying diagnostic, procedural, or prescription codes, participants without a relevant code were treated as not having the condition rather than as having missing data. Demographic covariates were drawn from self-reported enrollment data and were largely complete. Because All of Us enrolls volunteers rather than a probability-based sample, survey sampling weights were not applicable; group-level estimates were instead derived using marginal standardization. All data preprocessing and statistical analysis was done using RStudio Cloud Environment with R version 4.4.0.

## 3. Results

### 3.1. Descriptive Statistics (see Table 1)

We identified 370,722 adult participants with EHR data during the pre-defined study period. because the analytic cohort was drawn from existing health records, there were no recruitment stages and thus no participation-based attrition. 225,242 (60.8%) participants were female, 205,549 (55.4%) were White, 66,676 (18.0%) were Hispanic, and the median age was 55.1 years [Q1=39.1, Q3=66.5]. 49,140 (13.2%) people in the sample had at least 1 disability: 20,040 (5.4%) had 1+ sensory disabilities, 14,573 (3.9%) had 1+physical disabilities, 6,647 (1.8%) had 1+ cognitive disabilities, and 7,880 (2.1%) had more than one disability subtype. Chronic pain was observed in 17,739 (36.1%) of PWD and 38,675 (12.0%) of PWOD. 7328 (2.0%) of the sample had an OUD diagnosis, including 5,161 (1.6%) PWOD and 2,167 (4.4%) PWD. Of those, 2826 (38.6%) had received MOUD, 3702 (50.5%) had a chronic pain diagnosis, and 2892 (39.5%) had a remission diagnosis.

**Table 1:**
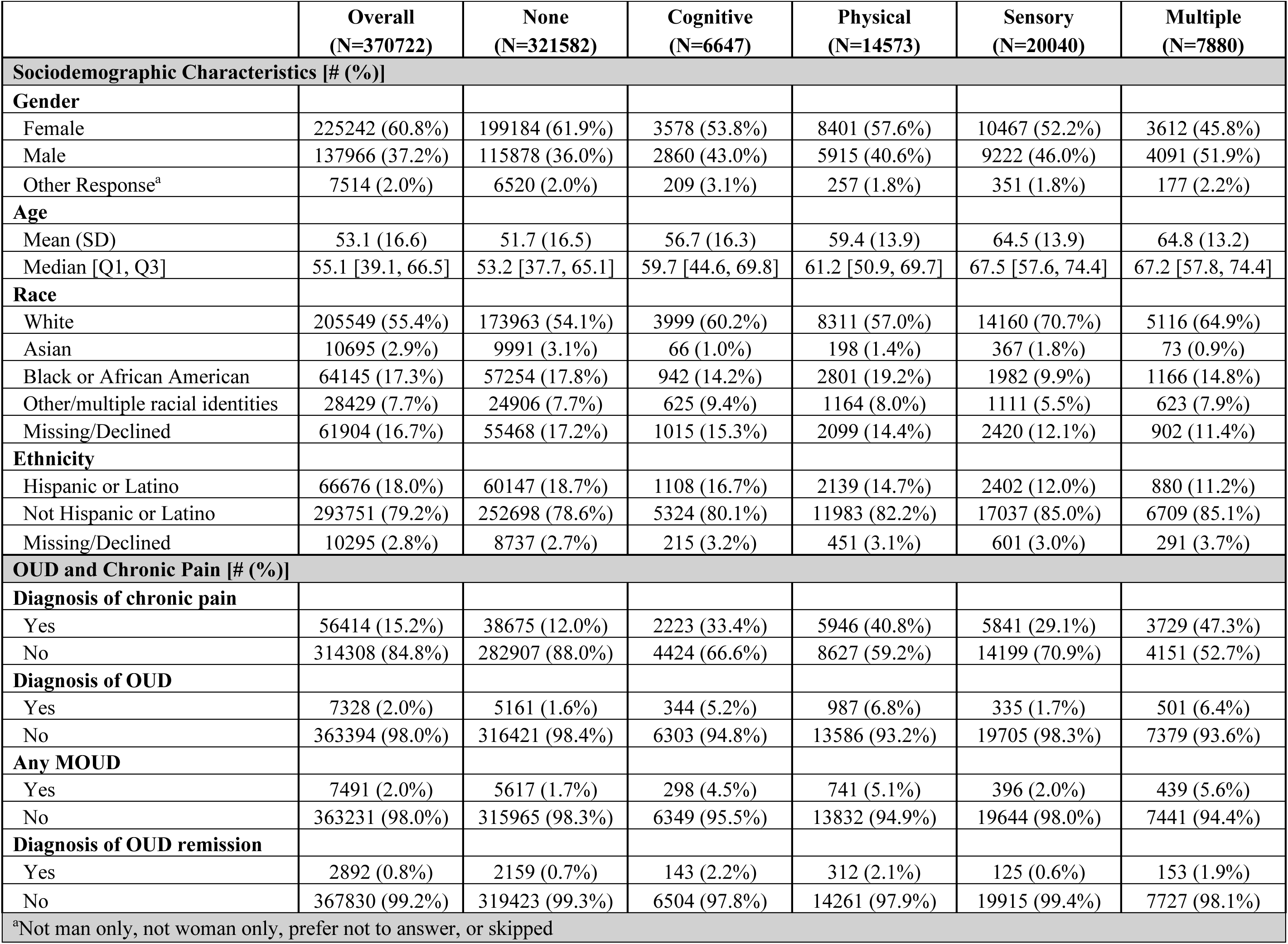
Sociodemographic and clinical characteristics of *All of Us s*ample.

### 3.2. Associations between disability, pain and OUD

After controlling for demographics and chronic pain, people with physical disabilities had 174% higher adjusted odds of an OUD diagnosis (aOR: 2.74, 95% CI: 2.54-2.95), people with multiple disability subtypes had 143% higher adjusted odds (2.43, 2.19-2.68), people with cognitive disabilities had 119% higher adjusted odds (2.19, 1.94-2.45), and people with sensory disabilities had 13% lower adjusted odds (0.87, 0.78-0.98), compared to PWOD (see **Table 2**). Participants with a chronic pain diagnosis had 497% higher adjusted odds of being diagnosed with OUD (5.97, 5.67-6.28). Chronic pain had a main effect on the predicted probability of OUD diagnosis across all subjects (see **Supplemental Table 2**). There was a statistically significant interaction between chronic pain and cognitive disability on the predicted probability of OUD diagnosis, compared to PWOD (see **Figure 1a** and **Supplemental Table 3**).

**Figure 1:**
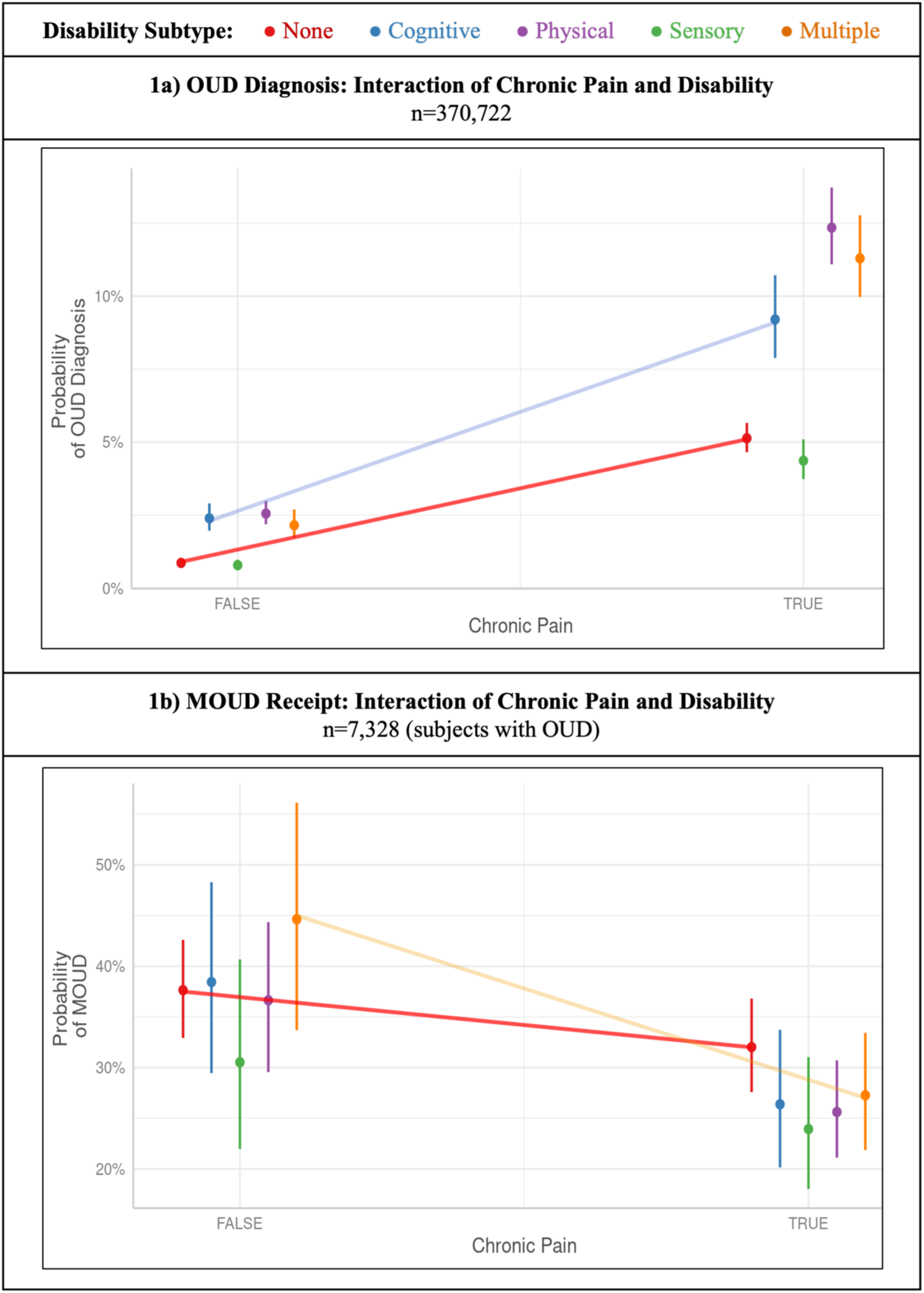
Interaction of disability subtype with chronic pain in predicting OUD diagnosis and MOUD receipt. Data points represent predicted probabilities calculated at the mean age (50.71 years for the whole sample; 53.08 years for the OUD diagnosis population) and weighted averages across categorical variables (gender, race, ethnicity, and chronic pain/MOUD when not the primary outcome). Error bars represent 95% confidence intervals. To highlight statistically significant effect modification, trend lines are superimposed for the reference group (No Disability) and disability subtypes exhibiting a significant interaction coefficient (p<0.05) relative to the reference group.

**Table 2:** Associations between OUD-related outcome and disability type, chronic pain, and MOUD receipt.

|  | OUD diagnosis |  | MOUD receipt |  | OUD remission |  |
| --- | --- | --- | --- | --- | --- | --- |
| *Predictors* | *OR (CI)* | *aOR<sup>a</sup> (CI)* | *OR (CI)* | *aOR<sup>a</sup> (CI)* | *OR (CI)* | *aOR<sup>b</sup> (CI)* |
| **Disability subtype** |  |  |  |  |  |  |
| **None** | (ref) | (ref) | (ref) | (ref) | (ref) | (ref) |
| **Cognitive** | **3.35**<br>(2.99 – 3.74) | **2.19**<br>(1.94-2.45) | **0.77**<br>(0.61 – 0.96) | 0.87<br>(0.69 – 1.10) | 0.99<br>(0.79 – 1.23) | **1.28**<br>(1.01 – 1.61) |
| **Physical** | **4.45**<br>(4.15 – 4.78) | **2.74**<br>(2.54-2.95) | **0.65**<br>(0.56 – 0.75) | **0.81**<br>(0.69 – 0.94) | **0.64**<br>(0.56 – 0.74) | 0.93<br>(0.80 – 1.09) |
| **Sensory** | 1.04<br>(0.93 – 1.16) | **0.87**<br>(0.78-0.98) | **0.56**<br>(0.44 – 0.71) | **0.7**<br>(0.54 – 0.90) | 0.83<br>(0.66 – 1.04) | 1.26<br>(0.99 – 1.61) |
| **Multiple** | **4.16**<br>(3.78 – 4.57) | **2.43**<br>(2.19 – 2.68) | **0.68**<br>(0.56 – 0.83) | 0.91<br>(0.74 – 1.11) | **0.61**<br>(0.50 – 0.74) | 0.97<br>(0.78 – 1.20) |
| **OUD-related variables** |  |  |  |  |  |  |
| **Chronic Pain** |  | **5.97**<br>(5.67 – 6.28) |  | **0.73**<br>(0.65 – 0.81) |  | **0.57**<br>(0.52 – 0.64) |
| **MOUD receipt** |  |  |  |  |  | **2.11**<br>(1.91 – 2.33) |
| <sup>a</sup>Controlling for demographic covariates (gender, age, race, and ethnicity) and chronic pain;<br><sup>b</sup>Controlling for demographic covariates, chronic pain, and MOUD<br>**Bold** values indicate significance at the p < 0.05 level |  |  |  |  |  |  |

### 3.3. Associations between disability, pain, and MOUD

Of participants with OUD diagnosis (n=7,328), people with sensory disabilities had 30% lower adjusted odds (aOR: 0.7, 95% CI: 0.54-0.90) and people with physical disabilities had 19% lower adjusted odds (0.81, 0.69-0.94) of receiving MOUD than PWOD after controlling for demographics and chronic pain. The adjusted odds of MOUD for people with cognitive (0.87, 0.69-1.10) and multiple disability subtypes (0.91, 0.74-1.11) were not significantly different than for PWOD (see **Table 2**). Across disability status, chronic pain was associated with lower adjusted odds of receiving MOUD (0.73, 0.65-0.81). Chronic pain was associated with significantly lower predicted probability of receiving MOUD within the no disability, physical disability, and multiple disability subtypes, but was not significant within the cognitive disability or sensory disability subtypes (see **Supplemental Table 2**). The predicted probability of MOUD was greatest for people with multiple disability subtypes and no chronic pain (0.45, 0.34-0.56), and lowest for people with sensory disability and chronic pain (0.24, 0.18-0.31). There was a significant interaction between multiple disability subtypes and chronic pain status when calculating the predicted probability of MOUD receipt (see **Figure 1b** and **Supplemental Table 3**).

### 3.4. Associations between disability, pain, and OUD remission

Among people with OUD (n=7,328), the adjusted odds of OUD remission for people with physical disability (aOR: 0.93, 95% CI: 0.80-1.09), sensory disability (1.26, 0.99-1.61) and multiple disability subtypes (0.97, 0.78-1.20) were not significantly different than for PWOD when controlling for demographics, chronic pain, and MOUD, though people with cognitive disabilities showed a modest increase in adjusted odds of remission (1.28, 1.01-1.61). Chronic pain was associated with decreased adjusted odds of OUD remission across disability status (0.57, 0.52-0.64) (see **Table 2**). Within those with multiple disabilities, the presence of chronic pain was associated with a lower predicted probability of OUD remission (0.28, 0.22-0.33) compared to those without chronic pain (0.48, 0.37-0.60) (see **Supplemental Table 2**). Those most likely to be diagnosed with remission had a sensory disability and no chronic pain (0.54, 0.43-0.65), while those with physical disability and chronic pain diagnosis were least likely (0.27, 0.22-0.32). The interaction of chronic pain and disability on predicted OUD remission probability was significant only for those with physical disability when compared with PWOD (see **Figure 2a** and **Supplemental Table 3**).

**Figure 2:**
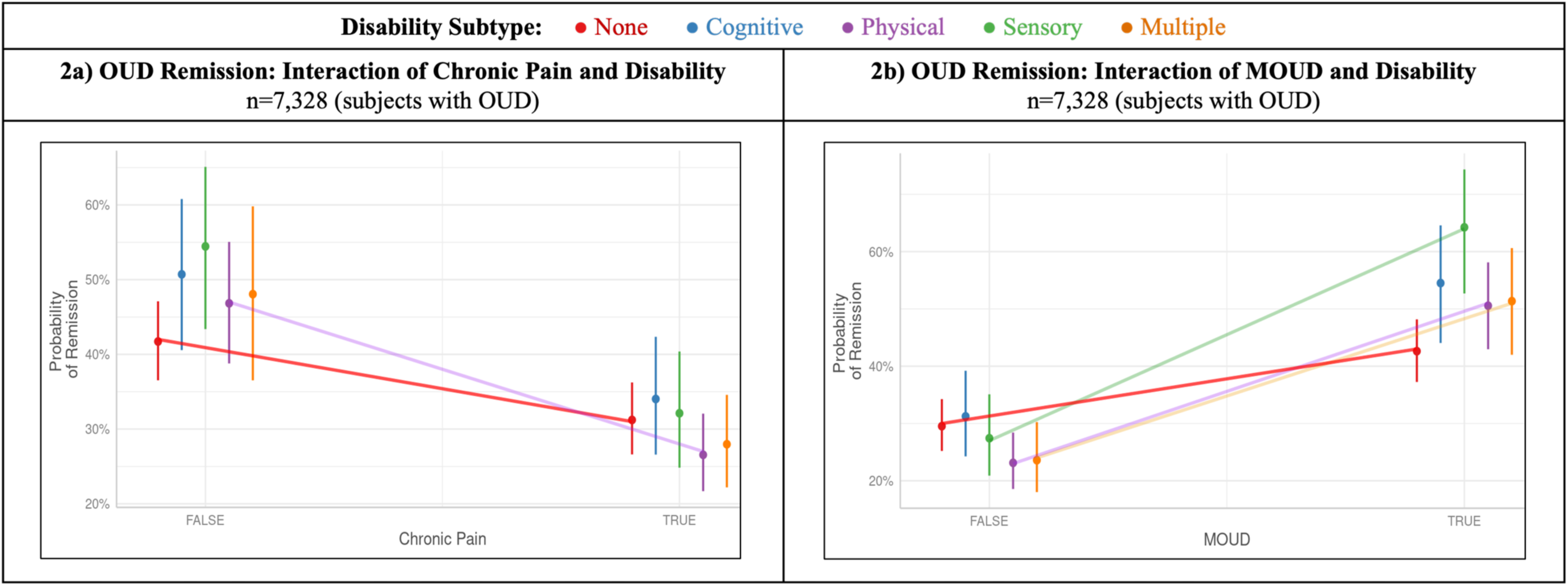
Interaction of disability subtype with chronic pain or MOUD receipt in predicting OUD remission. Data points represent predicted probabilities calculated at the mean age (53.08 years for the OUD diagnosis population) and weighted averages across categorical variables (gender, race, ethnicity, and chronic pain/MOUD when not the primary outcome). Error bars represent 95% confidence intervals. To highlight statistically significant effect modification, trend lines are superimposed for the reference group (No Disability) and disability subtypes exhibiting a significant interaction coefficient (p<0.05) relative to the reference group.

### 3.5. Associations between disability, MOUD receipt, and OUD remission

Across disability status, MOUD was associated with increased adjusted odds of OUD remission (aOR=2.11, 95% CI=1.91-2.33) (see **Table 2**). Patients who received MOUD also had a significantly increased likelihood of OUD remission within all disability subtypes (see **Supplemental Table 2**). The interaction between MOUD receipt and disability on OUD remission was significant for sensory, physical, and multiple disability subtypes (see **Figure 2b** and **Supplemental Table 3**). People with sensory disability who had received MOUD were most likely to be diagnosed with remission (0.64, 0.53-0.74), while people with physical disability who had not received MOUD were least likely (0.24, 0.19-0.28).

## 4. Discussion

In the *All of Us* sample, PWD of all subtypes exhibited a significantly higher predicted probability of OUD when controlling for demographic covariates, though those with physical and sensory disabilities were less likely than PWOD to receive MOUD. In the presence of chronic pain, those with cognitive, physical, or multiple disabilities showed a significantly higher predicted probability of being diagnosed with OUD, while the predicted probability of receiving MOUD was significantly lower for those with physical disabilities. Interaction terms revealed how chronic pain and receiving MOUD were differentially associated with OUD outcomes by disability subtype, compared to PWOD. Our findings highlight the significant disparities in OUD-related outcomes among PWD and further exhibit how the presence of chronic pain can further widen the substance use care gap across disability status.

Having any disability was associated with a greater likelihood of OUD after adjusting for demographic covariates. While those with physical, cognitive, and multiple disability subtypes exhibited a higher predicted probability of OUD, those with sensory disabilities showed a lower predicted probability. This pattern aligns with existing literature where OUD rates are higher in most disability subtypes, though people with hearing or vision loss show similar or lower rates of OUD and overdose.^4,5^ The association between chronic pain and higher predicted probability of OUD diagnosis was significantly stronger for those with cognitive disabilities compared to PWOD. However, given that cross-sectional data constrain causal inference, it is also possible that opioid use, further confounded by chronic pain, can result in cognitive disability.^28^ Altogether, the co-occurrence of disability, chronic pain, and OUD is not infrequent, thus highlighting the need for co-location of interdisciplinary teams specialized in OUD management for patients with complex needs.

Receipt of MOUD varied across disability subtypes, where those with physical and sensory disabilities had a significantly lower likelihood of receiving MOUD, potentially reflecting barriers related to access, stigma, or disjointed care. Although both OUD diagnoses and chronic pain^11^ are independent indications for MOUD, chronic pain was associated with a lower likelihood of MOUD receipt across the entire sample, especially among those with physical and multiple disabilities. The interaction between chronic pain and disability on MOUD receipt was largest among individuals with multiple disability subtypes, compared to PWOD, indicating that clinical complexity and competing pain management needs may reduce the likelihood of MOUD initiation in this vulnerable population. Better integration of OUD treatment and pain management through coordinated care models and targeted provider education may help reduce disparities in MOUD access among people with disabilities.^2,29^

There is minimal existing quantitative literature regarding OUD remission patterns in PWD,^30^ although qualitative studies do highlight how substance use treatment facilities are often inaccessible, and that stigma or under-education of providers regarding disability-inclusive care can preclude successful OUD recovery.^31^ Among our sample, the likelihood of OUD remission appeared lower in participants with physical or multiple disabilities than in PWOD, though these associations were not significant in the adjusted model. This trend implies a confounding relationship between chronic pain and intersectional identities, further complicating the likelihood of OUD remission and underscoring the importance of culturally competent, integrated care models that do not treat pain and OUD as competing priorities.^14^ Interestingly, after controlling for demographic covariates and chronic pain, people with cognitive disabilities surprisingly showed a significantly higher probability of OUD remission, compared to PWOD, potentially reflecting the protective role of caregiver oversight in treatment adherence and medication management.^32^

Chronic pain was associated with a decreased likelihood of OUD remission across all participants, even after adjusting for MOUD, suggesting that persistent pain may worsen psychological burden and sustain neurobiological pathways linked to opioid dependence.^14^ The predicted probability of OUD remission in the multiple disabilities group was significantly different for people with and without chronic pain, likely reflecting increased utilization of healthcare. Chronic pain was associated with a more significant decline in the predicted probability of OUD remission for those with physical disabilities, compared to PWOD, potentially reflecting the degree to which pain is intertwined with the disability itself.

Conversely, MOUD was consistently associated with an increased likelihood of remission across all participants, with predicted probabilities differing for all disability subtypes compared with PWOD. The interaction effects showed this relationship was significant among people with multiple, physical, and sensory disability subtypes, compared to PWOD. These gains are particularly noteworthy given that PWD often began with lower baseline remission probabilities. While the relative strength of association varied, MOUD remained a critical driver of recovery, regardless of disability status. Importantly, these results reinforce the need to ensure equitable access to MOUD for PWD, since the benefits appear both consistent and clinically meaningful.

## 5. Limitations

While our study offers valuable insights, the cross-sectional nature of the data precludes causal inference regarding the relationships between disability, chronic pain, and OUD outcomes. Future studies could incorporate the recorded dates of diagnostic and procedural codes to better understand longitudinal pathways between disability, chronic pain, OUD, MOUD access, and OUD remission. The study was conducted on a diverse national cohort, though not a nationally *representative* one; this has strengths for improving equity and representation in research, though the results may not be generalizable across all populations.

Disability and chronic pain were ascertained using diagnostic and procedural codes extracted from EHRs, following prior studies.^19–23^ Prior investigation within the *All of Us* Research Program suggested that EHR-derived measures underestimated the prevalence of vision and cognitive disabilities,^33^ raising the possibility that some participants were erroneously miscoded as PWOD. This aligns with our 13.3% prevalence of disability in the cohort, in contrast with the 28.7% disability rate in the U.S. adult population.^1^ Although our study design enhanced reproducibility and interpretability, the possibility remains that some disabilities were under-documented or missed, thus underestimating the true prevalence of these comorbidities, and potentially obscuring heterogeneity within categories. In addition, analyses primarily compared each disability subtype to PWOD rather than directly to each other, limiting inference about differences across subtypes. Future studies should integrate clinical and self-reported disability measures to further refine disability subtype classification and to understand between-group differences. Further, in stigmatized conditions such as OUD, it is likely that misuse, MOUD, and remission are under-documented in clinical records.^34^

## 6. Conclusions

Using a large, diverse, national cohort, this study yields quantitative evidence that disability and chronic pain are independently and synergistically associated with poorer OUD-related outcomes, and is among the first to quantify MOUD receipt rates and OUD remission metrics in PWD. Our analyses revealed that many PWD face higher odds of OUD, reduced MOUD access, and lower remission rates, with chronic pain significantly amplifying these disparities. Despite these challenges, MOUD remained a powerful driver of remission, underscoring the urgency of ensuring equitable substance use treatment access for PWD. Providers must be educated about the unique challenges faced by PWD to mitigate bias and improve clinical decision-making, and policies should incentivize health services accessibility, as well as efforts to enhance care coordination across pain management and addiction services.

## Data Availability

All data produced in the present study are available upon reasonable request to the authors

## Acknowledgements

We gratefully acknowledge All of Us participants for their contributions, without whom this research would not have been possible. We also thank the National Institutes of Health’s All of Us Research Program for making available the participant data in this study.

