## Supplemental Material for "Disability, Chronic Pain, and Opioid Use Disorder Treatment Disparities: A National Cohort Analysis from the *All of Us* Research Program"

Contents:

1) **Supplemental Table 1 (ST1):** Identification of disabilities and opioid use disorder-related diagnoses using medical codes
2) **Supplemental Table 2 (ST2):** Predicted probabilities of OUD-related outcomes by disability subtype
3) **Supplemental Table 3 (ST3):** Adjusted odds ratios for interaction effects between disability subtype, chronic pain, and MOUD receipt

| **ST1: Identification of disabilities and opioid use disorder-related diagnoses using medical codes** | | | | | |
| --- | --- | --- | --- | --- | --- |
| **Diagnosis Type** | | **ICD9** | **ICD10** | **CPT** | **HCPCS** |
| **Disability** | **Cognitive** | 317-319, 299.00, 299.80 | F70-F79, F84 | 97151-97158 |  |
|  | **Physical** | 343-344, 780.72, 799.3, V46, V49.6, V49.89, V62.89 | G80-83, R53.2, Z73.6, Z74, Z89, Z99 | 97535, 97542, 97761 | E0700, E0935, E0138, E1161, E1232, L5000-L9000, S9123-S9124 |
|  | **Sensory** | 20.96, 369, 389 | H54, H90-H91, Z96.2 |  |  |
| **Opioid Use Disorder (OUD)** | **OUD Diagnosis** | 304.0, 304.7, 305.5 | F11 |  |  |
|  | **OUD Remission** | 304.03, 304.73, 305.53 | F11.X1 |  |  |
| **Pain** | **Chronic Pain** | Z79.891, 33.82, 338.4 | G89.2, G89.4 |  |  |

ICD: International Classification of Diseases; ICD9 used 1979-9/30/2015; ICD10 used 10/1/2015-12/30/2021; CPT: Current Procedural Terminology; HCPCS: Healthcare Common Procedure Coding System

| **ST2: Predicted probabilities of OUD-related outcomes by disability subtype** | | | | | |
| --- | --- | --- | --- | --- | --- |
|  | **No Disability** | **Cognitive Disability** | **Physical Disability** | **Sensory Disability** | **Multiple Disabilities** |
| **OUD diagnosis [aOR (95% CI)]** | | | | | |
| **+ Chronic Pain** | 0.05 (0.05 – 0.06)^a^ | 0.09 (0.08 – 0.11)^b^ | 0.12 (0.11 – 0.14)^c^ | 0.04 (0.04 – 0.05)^d^ | 0.11 (0.10 – 0.13)^e^ |
| **- Chronic Pain** | 0.01 (0.01 – 0.01)^a^ | 0.02 (0.02 – 0.03)^b^ | 0.03 (0.02 – 0.03)^c^ | 0.01 (0.01 – 0.01)^d^ | 0.02 (0.02 – 0.03)^e^ |
| **MOUD receipt among those with OUD [aOR (95% CI)]** | | | | | |
| **+ Chronic Pain** | 0.32 (0.28 – 0.37)^f^ | 0.26 (0.20 – 0.34) | 0.26 (0.21 – 0.31)^g^ | 0.24 (0.18 – 0.31) | 0.27 (0.22 – 0.33)^h^ |
| **- Chronic Pain** | 0.38 (0.33 – 0.43)^f^ | 0.38 (0.29 – 0.48) | 0.37 (0.30 – 0.44)^g^ | 0.31 (0.22 – 0.41) | 0.45 (0.34 – 0.56)^h^ |
| **OUD remission among those with OUD [aOR (95% CI)]** | | | | | |
| **+ Chronic Pain** | 0.31 (0.27 – 0.36) | 0.34 (0.27 – 0.42) | 0.27 (0.22 – 0.32) | 0.32 (0.25 – 0.40) | 0.28 (0.22 – 0.35)^i^ |
| **- Chronic Pain** | 0.42 (0.37 – 0.47) | 0.51 (0.41 – 0.61) | 0.47 (0.39 – 0.55) | 0.54 (0.43 – 0.65) | 0.48 (0.37 – 0.60)^i^ |
| **OUD remission among those with OUD [aOR (95% CI)]** | | | | | |
| **+ MOUD** | 0.43 (0.37 – 0.48)^j^ | 0.55 (0.44 – 0.65)^k^ | 0.51 (0.43 – 0.58)^l^ | 0.64 (0.53 – 0.74)^m^ | 0.51 (0.42 – 0.61)^n^ |
| **- MOUD** | 0.3 (0.25 – 0.34)^j^ | 0.31 (0.24 – 0.39)^k^ | 0.23 (0.19 – 0.28)^l^ | 0.27 (0.21 – 0.35)^m^ | 0.24 (0.18 – 0.30)^n^ |

Predicted probabilities are calculated for mean age and weighted averages across categorical variables (gender, race, ethnicity, MOUD, chronic pain). Predicted probability pairs with the same superscript (a-n) differed in post hoc pairwise comparisons, at p < 0.01

| **ST3: Adjusted odds ratios for interaction effects between disability subtype, chronic pain, and MOUD receipt [aOR (95% CI)]** | | | | |
| --- | --- | --- | --- | --- |
| **Disability Subtype** | **OUD Diagnosis:  Pain Interaction^a^** | **MOUD Receipt:**  **Pain Interaction^a^** | **OUD Remission:**  **Pain Interaction^a^** | **OUD Remission:**  **MOUD Interaction^b^** |
| **None** | Ref | Ref | Ref | Ref |
| **Cognitive** | **0.67 (0.53** – **0.84)** | 0.74 (0.46 – 1.18) | 0.79 (0.49 – 1.27) | 1.49 (0.92 – 2.42) |
| **Physical** | 0.86 (0.74 – 1.02) | 0.76 (0.55 – 1.06) | **0.65 (0.46 – 0.90)** | **1.92 (1.40 – 2.63)** |
| **Sensory** | 0.93 (0.73 – 1.18) | 0.92 (0.55 – 1.54) | 0.62 (0.37 – 1.03) | **2.68 (1.58 – 4.62)** |
| **Multiple** | 0.93 (0.73 – 1.19) | **0.60 (0.37 – 0.97)** | 0.66 (0.40 – 1.09) | **1.93 (1.26 – 2.97)** |

OUD: opioid use disorder; MOUD: medications for OUD; aOR: adjusted odds ratio; CI: confidence interval

**Bold** values indicate statistically significant interaction effects (p<0.05)

^a^Models adjusted for sociodemographic covariates (gender, race, ethnicity, age at beginning of study period)

^b^Models adjusted for sociodemographic covariates and presence of chronic pain

**Note:** The interaction aOR represents the change in the odds of the outcome associated with the primary predictor (chronic pain or MOUD receipt) for a specific disability subtype relative to the change observed in the reference group (No Disability).
